# PRIMARY CENTRAL NERVOUS SYSTEM VASCULITIS WITH INTRACRANIAL ANEURYSM

**DOI:** 10.1101/2024.01.22.24301637

**Authors:** Carlo Salvarani, Robert D. Brown, Teresa J. H. Christianson, John Huston, Caterina Giannini, Gene G. Hunder

## Abstract

**Background:** Unruptured intracranial aneurysms (UIAs) are rarely reported in primary central nervous system vasculitis (PCNSV). In this study we described the clinical findings, response to therapy, and outcomes of UIA in a large cohort of PCNSV patients.

**Methods:** We retrospectively studied 216 consecutive patients with PCNSV, selected by predetermined diagnostic criteria, who were seen during a 40-year period. UIAs were identified on cerebral angiography. The clinical, laboratory, radiologic and pathologic findings, management, and outcomes of patients with UIA were described and compared with those without UIA.

**Results:** 12/216 (5.5%) PCNSV patients had at least one UIA. In the only positive patient biopsy showed necrotizing vasculitis. Eleven patients had evidence of UIA on angiogram at diagnosis. One patient developed an aneurysm during the follow-up associated with a worsening of vasculitic radiological findings. The most common presenting symptom for PCNSV in the setting of UIA was headache (67%), followed by persistent neurologic deficit or stroke (50%). Most patients with UIA presented with multiple cerebral infarcts on MRI (67%), one patient had subarachnoid hemorrhage, and one left parieto-occipital intracerebral hematoma, both unrelated to the aneurysm. Black blood imaging was performed in 4 patients and 2 showed segmental circumferential mural enhancement involving multiple vessels. Two patients had 2 UIAs, while the other 10 had 1. The most frequent UIA location was internal carotid artery (50%), followed by anterior cerebral artery (21%). Ten of the UIAs were < 5 mm in diameter, and 3 were 5-7 mm in diameter; the size was not available for one. All UIAs were unchanged in size and configuration during follow-up and no new aneurysms were detected. Compared to the 204 patients with PCNSV without a UIA, no significant clinical differences were observed, except for a reduced disability at last follow-up (p = 0.038).

**Conclusions:** UIAs uncommonly occur in PCNSV.

---

Primary central nervous system vasculitis (PCNSV) is an uncommon and poorly understood vasculitis restricted to brain and spinal cord (1–4). Calabrese and Mallek defined in 1988 the diagnostic criteria that give equal weight to angiography and brain biopsy (5). Although characterized by a limited specificity, angiographic findings suggesting a high probability of vasculitis include areas of smooth-wall segmental narrowing or dilation and occlusions that affect multiple cerebral arteries without proximal vessel changes of atherosclerosis or other possible causes (1, 3, 4). Intracranial aneurysms are rarely reported in PCNSV (7–10).

In this study, we evaluated all radiological examinations of 216 consecutive patients with PCNSV evaluated at the Mayo Clinic from 1983 to 2022. The aim was to determine and report the characteristics of all PCNSV cases who had radiological evidence of at least one unruptured intracranial aneurysm (UIA) at the time of PCNSV diagnosis or during follow-up. We also compared PCNSV patients with and without a co-occurring UIA.

## METHODS

### Identification of patients

The updated cohort of 216 consecutive patients with PCNSV evaluated at the Mayo Clinic over a 40-year period from 1983 to 2023 was utilized for the study. The same predefined diagnostic criteria were used to identify all patients (4, 11). All biopsy specimens were reviewed by the same neuropathologist (CG) and a neuroradiologist reviewed all cerebral angiograms and other imaging studies. Angiogram findings were divided in two groups: large/proximal artery (intracranial internal carotid and vertebral arteries, basilar artery, and proximal anterior, middle, and posterior cerebral arteries) and small artery involvement (second division branches or smaller). Patients with identification of UIA at diagnosis or during follow-up were identified. Available cerebral angiograms performed at the beginning or before the beginning of PCNSV clinical manifestations, at PCNSV diagnosis and during follow-up were revaluated. Radiological follow-up was evaluated from the first evidence of aneurysm by imaging to the last angiography, magnetic resonance angiography (MRA), and/or computed tomography angiography (CTA) available during the follow-up. The median duration of the follow-up for the total 216 patients was 22.2 months (range 0 to 337 months).

### Standard Protocol Approvals, Registrations, and Patient Consents

The study was approved by the Mayo Clinic Institutional Review Board. Written informed patient consent to perform this study was received by all patients.

### Clinical data collection

Detailed information was recorded from Mayo Clinic medical records, at diagnosis and during follow-up. This included clinical manifestations, other medical conditions, laboratory investigations, radiological imaging, results of CNS biopsy or autopsy, management, relapses, follow-up functional status and cause of death (4,11). To determine the effect of treatment, we used the treating physician’s global opinion about the response to therapy as recorded in the detailed medical record. A neurologist examined all patients at PCNSV diagnosis and on subsequent visits including the last visit or death.

Relapse was defined as a recurrence or worsening of symptoms or progression of existing or new lesions on subsequent magnetic resonance imaging (MRI) examinations while the patient received no medication or a stable dose. A diagnosis of relapse required an increase in therapy. The degree of disability at presentation and at the last visit was defined through a review of the detailed clinical data in the medical record and was categorized by the modified Rankin Scale (12).

### Statistical analysis

We used a 2-sided 2-sample t test to compare numerical parameters or a Wilcoxon rank-sum test when the distributions were skewed. Fisher exact test was used for categorical parameters. Rankin scores were dichotomized into 0-3 and 4-6 because the difference between these 2 categories was medically relevant.

Survival was estimated with the Kaplan-Meier method. Cox proportional hazards modeling was performed for age adjusted survival comparison. All p values were 2 sided; significance was defined at p < 0.05. The statistical analysis was performed with SAS version 9 (SAS Institute Inc, Cary, NC).

### Data Availability

Anonymized data not published within this article will be made available by request from any qualified investigator.

## RESULTS

This retrospective single-center cohort study included 216 patients diagnosed with PCNSV at Mayo Clinic between 1983 and 2022. Vasculitis was found on biopsy in 74 patients, while angiography alone was used to confirm the diagnosis in 142 patients.

Twelve of the 216 patients (5.5%) had at least one UIA on angiography at PCNSV diagnosis (11 patients) or after the diagnosis, during follow-up (1 patient). One of the 12 patients with UIA had a normal cerebral angiogram 10 months before the PCNSV diagnosis.

### Demographic and clinical features

Most patients were women (75%). The median time from onset of PCNSV manifestations to diagnosis was 0.7 months (range 0-13 months) (Table 1). The most common symptom at presentation was headache followed by persistent neurologic deficit or stroke. Other medical disorders were observed in 8 patients, including hypertension in 5 patients, asthma in 2, thyroid gland disorders in 2, and type 2 diabetes in 1. Six patients were current smokers.

**Table 1.** Findings at PCNSV diagnosis and outcomes comparing those with and without intracranial aneurysm*.

|  | Absence of intracranial aneurysm (n=204) | Presence of intracranial aneurysm (n=12) | P value |
| --- | --- | --- | --- |
| Male | 93 (45.6) | 3 (25) | 0.234 |
| Age at diagnosis, years | 49 (17-85) | 56.5 (35-73) |  |
| Time from symptom onset to PCNSV diagnosis, months | 1.6 (0.0-120.6) | 0.7 (0.0-13.0) |  |
| Clinical manifestations at presentation |  |  |  |
| Headache | 114 (55.9) | 8 (66.7) | 0.558 |
| Cognitive dysfunction | 108 (52.9) | 5 (41.7) | 0.556 |
| Persistent neurologic deficit or stroke | 91 (44.6) | 6 (50.0) | 0.771 |
| Hemiparesis | 86 (42.2) | 5 (41.7) | 1.000 |
| Aphasia | 48 (23.5) | 1 (8.3) | 0.306 |
| Transient ischemic attack | 53 (26.0) | 2 (16.7) | 0.735 |
| Ataxia | 35 (17.2) | 3 (25.0) | 0.447 |
| Seizures | 40 (19.6) | 2 (16.7) | 1.000 |
| Visual symptoms (any kind) | 68 (33.3) | 5 (41.7) | 0.545 |
| Visual field defect | 30 (14.7) | 4 (33.3) | 0.100 |
| Blurred vision or decreased visual acuity | 22 (10.8) | 0 | 0.616 |
| Intracranial hemorrhage | 17 (8.3) | 2 (16.7) | 0.285 |
| Paraparesis or quadriparesis | 9 (4.4) | 0 | 1.000 |
| Systemic manifestations<sup>§</sup> | 17 (8.3) | 1 (8.3) | 1.000 |
| Fever | 16 (7.8) | 1 (8.3) | 1.000 |
| ESR, mm/hour<sup>^</sup> | 10 (0-124) | 8.5 (1-61) |  |
| Modified Rankin Score at last follow-up& |  |  |  |
| 0-3 | 139 | 11 | 0.038 |
| 4-6 | 56 | 0 |  |
| No relapse during follow-up & | 129/192 (67.2) | 9/11 (81.8) | 0.508 |
| Therapy response& | 156/189 (82.5) | 11/11 (100.0) | 0.217 |

\*Continuous data are presented as median and range. Categorical data are presented as number and percentage of patients. PCNSV = primary central nervous system vasculitis. ESR = erythrocyte sedimentation rate. §Defined as the presence of at least 1 of the following: fatigue, anorexia, weight loss, or fever. ^ESR was available for 174 of the 204 patients without aneurysms and for all the 12 patients with aneurysms. &Only patients with a follow-up duration of at least 2 weeks were included.

No patients developed varicella zoster virus (VZV) infection 12 months before or 6 months after PCNSV diagnosis.

### Laboratory investigations

The results of cerebrospinal fluid (CSF) examinations and the value of erythrocyte sedimentation rate (ESR) are shown in Table 2. CSF examinations were abnormal in all 11 patients with UIA who underwent spinal tap. Venereal Disease Research Laboratory test, fungal and viral serologies, particularly anti-VZV immunoglobulin G and anti-herpes simplex virus (HSV)-1 antibodies, and VZV and HSV DNA were evaluated in 5 patients and were negative. Morphologic evaluation and/or immunocytochemical studies were evaluated in 4 patients and were negative.

**Table 2.** CSF and MRI results at PCNSV diagnosis comparing those with and without intracranial aneurysm*.

|  | Absence of intracranial aneurysm (n=204) | Presence of intracranial aneurysm (n=12) | P value |
| --- | --- | --- | --- |
| CSF |  |  |  |
| Protein > 45 mg/dl ^ | 123/159 (77.4) | 10/11 (90.9) | 0.459 |
| WBC count > 5 cells/mm<sup>3</sup> | 86/159 (54.1) | 6/11 (54.5) | 1.000 |
| Protein > 45 mg/dl or WBC count > 5 cells/mm<sup>3</sup> | 130/161 (80.7) | 11/11 (100.0) | 0.218 |
| Protein > 70 mg/dl or WBC count > 10 cells/mm<sup>3</sup> | 102/159 (64.2) | 5/11 (45.5) | 0.333 |
| Initial MRI findings<sup>§</sup> |  |  |  |
| Presence of infarct | 104/188 (55.3) | 8/11 (72.7) | 0.354 |
| Single infarct | 11/188 (5.9) | 1/11 (9.1) | 0.505 |
| Multiple infarcts, in the same hemisphere | 20/188 (10.6) | 3/11 (27.3) | 0.120 |
| Multiple infarcts bilateral | 73/188 (38.8) | 4/11 (36.4) | 1.000 |
| Gadolinium-enhancing lesions (intracerebral or meningeal) | 71/188 (37.8) | 5/11 (45.5) |  |
| Intracerebral gadolinium-enhanced lesions | 40/188 (21.3) | 2/11 (18.2) | 0.751 |
| Meningeal gadolinium-enhanced lesions | 38/188 (20.2) | 4/11 (36.4) | 0.249 |
| Intracerebral hemorrhage& | 14 /188 (7.4) | 1/12 (8.3) | 1.000 |
| Subarachnoid hemorrhage | 3/188 (1.6) | 1/11 (9.1) | 0.205 |

\*Categorical data are presented as number and percentage of patients. PCNSV = primary central nervous system vasculitis. ^The normal range of protein is 14-45 mg/dL. §Magnetic resonance imaging (MRI) was performed initially in 188 of the 204 patients without aneurysm and in 11 of the 12 patients with aneurysm. &Including also patients with evidence of intracerebral hemorrhage on CT scan. One patient with intracranial aneurysm had intracerebral hemorrhage at presentation, however the diagnosis was made using CT scan; CSF = cerebrospinal fluid. WBC = white blood cell.

ESR at diagnosis was normal in 11 of the 12 patients (92%) (median 8.5 mm/hour; range 1-61).

### Radiological imaging

Eleven patients (92%) had UIA on cerebral angiography at the time of PCNSV diagnosis (Table 3 and Figure 1). In one of these patients (case 3) the first angiogram was normal, performed at the beginning of neurological manifestations 10 months before PCNSV diagnosis. In the other patient (case 10), an aneurysm was detected during follow-up (8 months after PCNSV diagnosis) in association with the appearance of new infarctions. There was no evidence of UIA on initial angiography.

**Table 3.** Demographic, diagnostic and imaging findings of the 12 patients with PCNSV and intracranial aneurysm.

| Case | Age, y* | Angiogram/<br>cerebral biopsy<br>findings of<br>PCNSV | Arterial imaging | Intracranial aneurysm location and size, and<br>relationship between aneurysm and PCNSV<br>diagnoses | Other lesions at MRI/MRA when<br>aneurysms were detected |
| --- | --- | --- | --- | --- | --- |
| 1 | 65-74 | +/ND | angiography | - 2 mm aneurysm of the left supraclinoid ICA<br>- at PCNSV diagnosis | multiple infarcts |
| 2 | 35-44 | +/ND | angiography and<br>MRA | - 6.5X7 mm aneurysm arising along the distal<br>margin of left PICA origin<br>- at PCNSV diagnosis | Non-enhancing T2 signal abnormalities |
| 3 | 35-44 | +/ND | angiography | - 2-3 mm aneurysm involving a left<br>proximal frontopolar branch and 1-2 mm<br>aneurysm involving the right PCA<br>- at PCNSV diagnosis. However, a cerebral<br>angiogram performed 10 months before, at<br>the appearance of neurological manifestations,<br>was normal | multiple infarcts and foci of abnormal<br>contrast enhancement primarily involving<br>the cortex and the adjacent leptomeninges |
| 4 | 45-54 | +/ND | angiography, MRA | - 2 mm aneurysm involving the posterior<br>aspect of the right supraclinoid ICA and 2<br>mm aneurysm of the right posterior<br>cerebral artery near the P1-2 junction<br>- at PCNSV diagnosis | multiple infarcts |
| 5 | 45-54 | +/ND | angiography | - 1-2 mm aneurysm, right supraclinoid ICA. Also<br>had small pseudoaneurysm in the mid-cervical<br>right ICA.<br>- at PCNSV diagnosis | subarachnoid hemorrhage not thought to<br>be related to aneurysm |
| 6 | 55-64 | +/- | angiography | - aneurysm of the suprasellar right ICA<br>- at PCNSV diagnosis | left parieto-occipital intracerebral<br>hematoma not thought to be related to<br>aneurysm |
| 7 | 65-74 | +/ND | angiography and<br>MRA | - 1.5 x 2 mm aneurysm, left MCA<br>bifurcation<br>- at PCNSV diagnosis | gadolinium enhancing cerebral lesion |
| 8 | 55-64 | +/ND | angiography and | - 5 mm aneurysm, origin of the left | multiple infarcts and leptomeningeal |
|  |  |  | MRA | ophthalmic artery<br>- at PCNSV diagnosis | enhancement |
| 9 | 35-44 | + / ND | angiography and MRA | - 5X6 mm aneurysm, left paraclinoid ICA<br>- at PCNSV diagnosis | multiple infarcts.<br>HR contrast-enhanced black blood imaging (6 months after starting PDN and MMF): no abnormal mural enhancement, particularly of the wall of the aneurysm |
| 10 | 55-64 | + / ND | MRA | - 2 mm aneurysm, right posterior communicating artery origin<br>- during follow-up. It was absent on angiography and MRA performed at diagnosis and was present on 3 MRAs performed 8 months, 23 months, and 31 months after the diagnosis. At the time of detection worsening of the brain MRI with appearance of new infarctions. | Single infarct at diagnosis. When cerebral aneurysm was detected: multiple infarcts. HR contrast-enhanced black blood imaging: vessel wall enhancement along the right supraclinoid/terminal ICA, also extending more proximally along the cavernous and petrous segments, and along the proximal basilar artery and the proximal right PCA |
| 11 | 55-64 | + / ND | angiography, MRA, CTA | - 3X2 mm aneurysm arising from the left A1/ anterior communicating artery junction<br>- at PCNSV diagnosis | multiple infarcts.<br>HR contrast-enhanced black blood imaging: mural enhancement along the right MCA bifurcation region, and the left M1 and bifurcation regions |
| 12 | 65-74 | - / + | angiography, CTA and MRA | - 3 mm aneurysm arising from the anterior communicating artery<br>- at PCNSV diagnosis | leptomeningeal enhancement with multiple infarcts and myriad of punctate areas of micro- hemorrhage throughout both cerebral hemispheres.<br>HR contrast-enhanced black blood imaging (3 months after starting PDN and oral CYC): no mural enhancement |

\*age variable is replaced by a generalized version with intervals of 10 years. Abbreviations: PCNSV = primary central nervous system vasculitis; MRI = magnetic resonance imaging; MRA = magnetic resonance angiography; ND = not done; ICA = internal carotid artery; PICA = posterior inferior cerebellar artery; PCA = posterior cerebral artery; ICA = internal carotid artery; MCA = middle cerebral artery; HR = high resolution; PDN = prednisone; MMF = mycophenolate mofetil; CTA = computed tomography angiography; CYC = cyclophosphamide.

**Figure 1.**
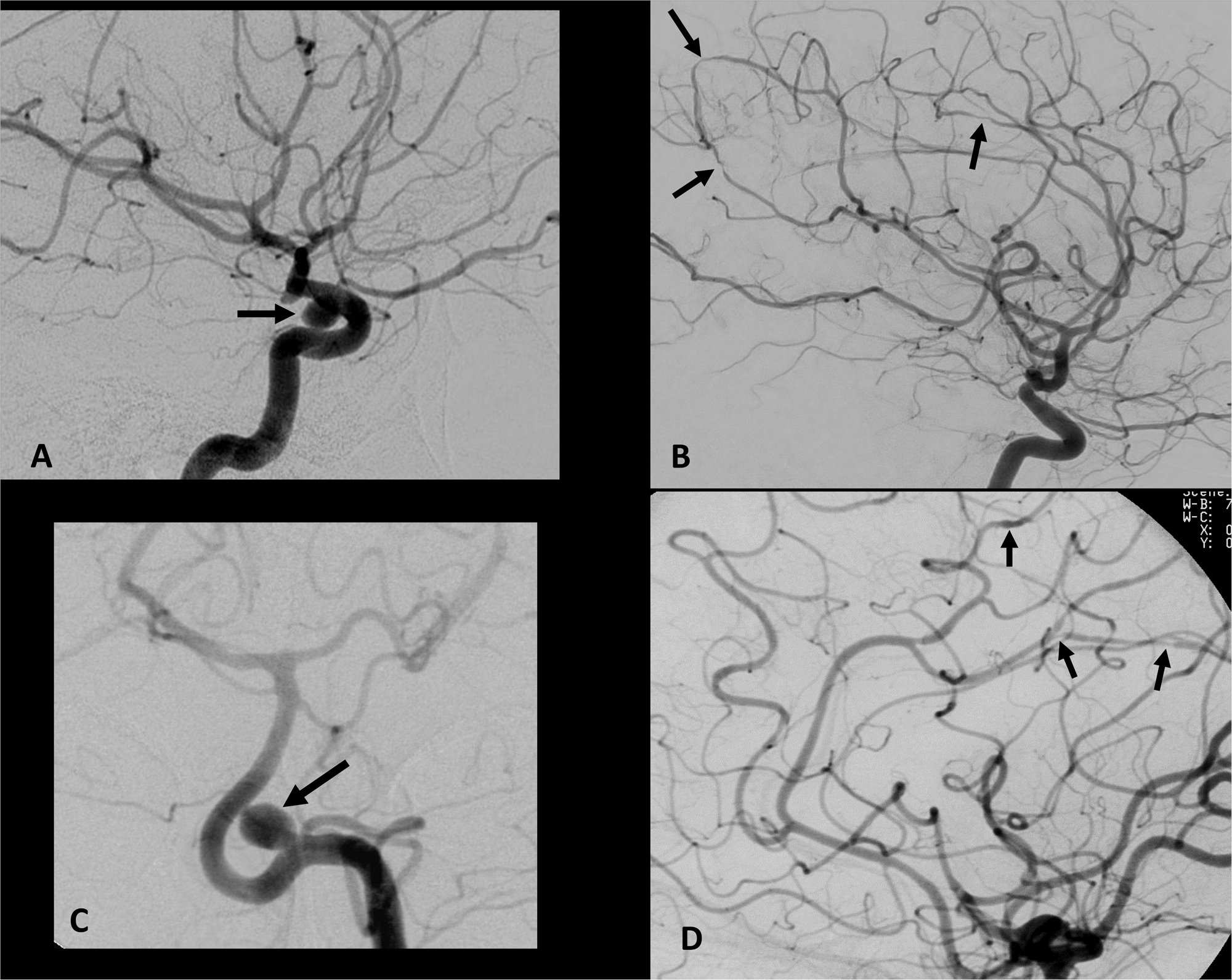
Cerebral angiograms demonstrating intracranial saccular aneurysms in the presence of PCNSV. Case 9: Left 6 × 5 mm left paraclinoid ICA aneurysm (arrow A). No evidence of the aneurysm following treatment with an intravascular pipeline device, however multiple focal zones of arterial narrowing remain (arrows B) at follow-up imaging. Case 2: Left 6.5 × 7 mm posterior inferior cerebellar artery aneurysm (arrow C). Left ICA injection demonstrates multiple focal zones of arterial narrowing and irregularity (arrows D).

**Figure 2.**
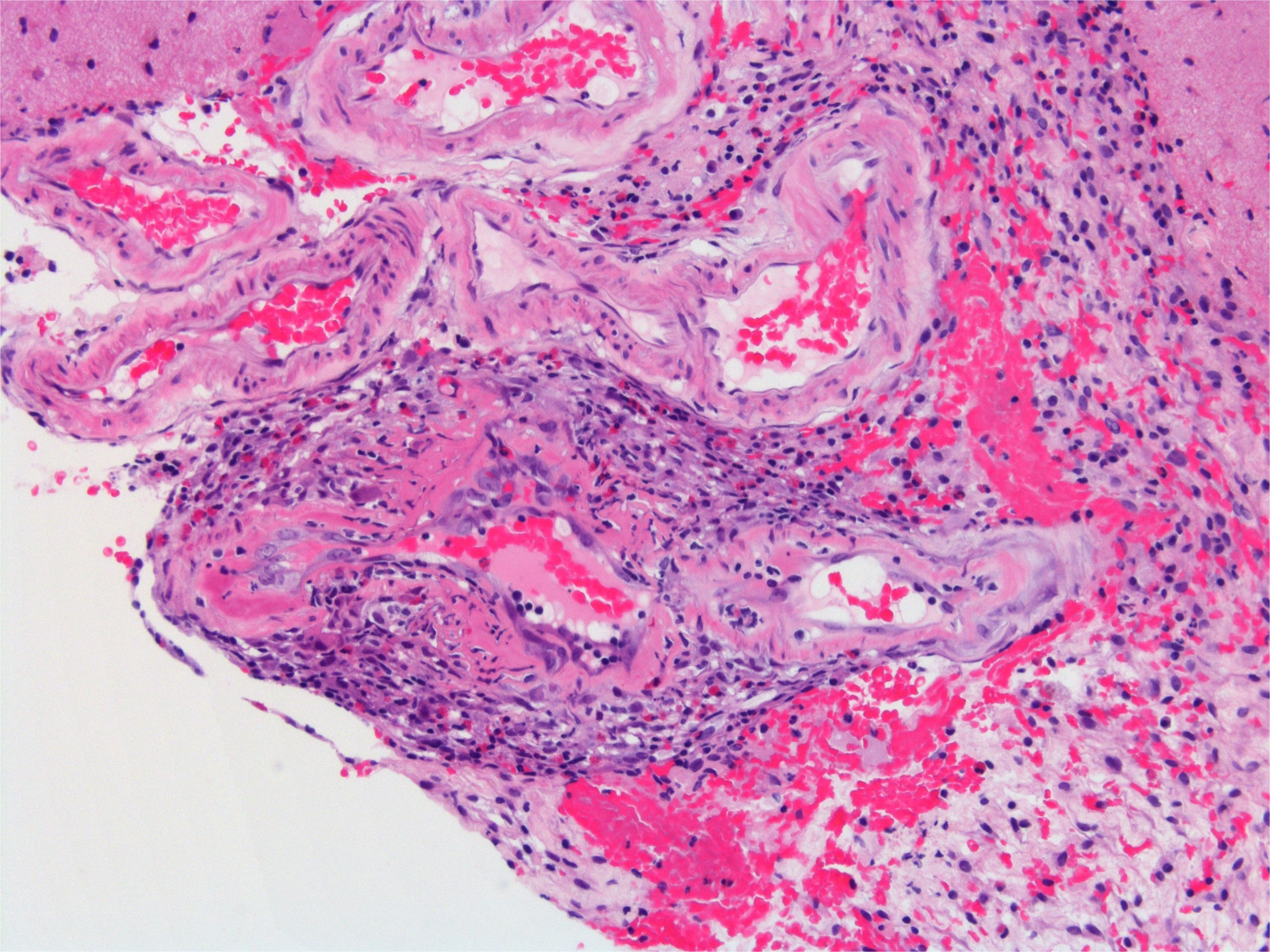
Pathologc findings of cerebral biopsy of case 12. Photomicrography showing transmural acute inflammation and segmental transmural fibrinoid necrosis involving a leptomeningeal small artery, without giant cells or granulomas. (Hematoxylin and eosin stained; original maginification X 200).

Table 2 also summarizes the neuroimaging at diagnosis. Cerebral infarctions were seen in 8 of the 12 patients who underwent MRI and/or CT scans at diagnosis; infarctions were multiple in 7 patients and bilateral in 4. Infarction was single at diagnosis in one patient (case 10), but she developed multiple infarcts during the follow-up. Parenchymal and/or leptomeningeal enhancing lesions were observed in 5 patients, subarachnoid hemorrhage (SAH) in 1 patient, and left parieto-occipital intracerebral hemorrhage (ICH) in 1 patient. The SAH and ICH were not felt to be caused by the intracranial aneurysm. In the last patient the diagnosis of ICH was made using CT-scan.

High-resolution contrast-enhanced black blood imaging was performed in 4 patients. Segmental circumferential mural enhancement involving multiple vessels was observed in 2 patients (cases 10 and 11), while there was no mural enhancement in the other two cases (cases 9 and 12). However, in these two last cases black blood imaging was obtained after 6 months and 3 months of immunosuppressant treatment, respectively (Table 3).

In 11 of the 12 patients PCNSV diagnosis was made on angiography alone, while one patient had negative angiography and cerebral biopsy showing vasculitis (case 12). Four patients had at least 2 angiograms. Multiple bilateral intracranial arterial stenoses were present on angiography in 10 patients, while one patient had multiple unilateral lesions. In 4 patients angiography showed involvement of both large/small arteries, while in the other 7 only small cerebral arteries were involved.

The UIA arterial location and diameter are found in Table 3. There were two UIAs in 2 patients and one UIA in the other 10. The arterial location of the 14 UIAs were as follows: 7 internal carotid artery (ICA), 3 anterior cerebral artery (ACA), 2 posterior cerebral artery (PCA), 1 middle cerebral artery (MCA), and 1 posterior inferior cerebellar artery (PICA). Ten of the UIAs were < 5 mm in diameter, and 3 were 5-7 mm in diameter. In one patient the aneurysm size was not available.

### Biopsy results

Two of 12 patients underwent brain biopsy. Histological evidence of necrotizing vasculitis was found in one patient (case 12). Amyloid angiopathy was also present, however, the amyloid deposition was not strictly associated with the vasculitic lesions. In the second patient the biopsy was negative (case 6).

### Treatment and outcome

All 12 patients received oral glucocorticoids. In 5 patients, prednisone was the only therapeutic agent used. In 5 patients, intravenous pulse methylprednisolone 1 gm/day for 3 to 9 days (median 5 days) was used before oral prednisone treatment was started. The median initial prednisone dose was 60 mg/day (range 40-80 mg/day). The median duration of oral prednisone therapy was 10 months (range 1-29 months).

Seven patients received immunosuppressants in addition to prednisone. Cyclophosphamide was used in 5 patients (4 received daily oral doses and 1 intravenous pulses). The median starting dose of oral cyclophosphamide was 100 mg/day, and the median therapy duration 3.5 months (range 1-10 months). One patient received intravenous pulse cyclophosphamide 800 mg/month for 6 months. Mycophenolate mofetil (2,000 mg/day for 23 months) and azathioprine (100 mg/day for 3 months) were used in the other two patients.

Two patients had a flare in PCNSV that led to a change in therapy. The first patient had a flare characterized by headache and worsening of MRI findings after 4 years of complete remission without therapy. He received intravenous pulse methylprednisolone followed by oral prednisone and restarted mycophenolate mofetil therapy. No follow-up information was reported. The second patient had two flares characterized by new subacute cerebral infarctions at MRI. She had the second flare during treatment with intravenous pulse cyclophosphamide started for the first flare, along with prednisone 20 mg/day. Cyclophosphamide was stopped and rituximab was started; oral prednisone was increased to 90 mg/day. Eighteen months later she was in remission and on treatment with prednisone 5 mg/day.

All 11 patients for which data were available responded to therapy and recovered with little or no residual disability. In these 11 patients, the median modified Rankin score at last visit was 1 (range 0-2). This was lower than at presentation (median 2.5; range 1-4). In one patient the therapy response was not evaluable. This patient started the therapy at diagnosis, but his follow-up was only 11 days and it was not possible to evaluate the treatment response.

Of the 192 patients without UIA with data on flares available, 63 (32.8%) had at least one flare, while of the 189 patients without UIA with data on treatment response available, 156 (82.5%) responded to therapy (Table 1).

### Aneurysm follow-up

Radiologic follow-up for the UIA was available in 10 patients, a median of 18.5 months (range 1-151 months) after initial assessment. No follow-up imaging was available in the other two. All UIAs were unchanged on follow-up imaging and no additional UIAs were detected. Patient 9 had a pipeline stenting of the aneurysm 24 months after diagnosis (Figure 1).

No patients had intracerebral or subarachnoid hemorrage during follow-up.

### Comparison of patients with and without intracranial aneurysm

The 12 patients with UIA were compared with the 204 without in Tables 1 and 2. No significant differences in demographic characteristics, spinal fluid results and neuroimaging findings at diagnosis were observed between the 2 groups. The frequencies of patients with relapsing disease and the response rate to treatment were similar.

The median duration of follow-up for the 12 patients with UIA was 35 months with a range from 12 days to 20 years. Patients with UIA had a statistically significant lower frequency of bad outcome (modified Rankin disability score ≥ 4) at last follow-up (p = 0.04). An increased survival trend for UIA patients was also observed: at the end of follow-up no patients with aneurysms died, compared to 46 (22.5%) of the 204 patients without UIA (p = 0.09).

## DISCUSSION

Unruptured intracranial aneurysms have been described uncommonly in PCNSV (7–10). In our series, UIA was detected at diagnosis or during follow-up in 12 of 216 patients (5.5%) with PCNSV. Findings from imaging studies suggest that the frequency of intracranial aneurysms in the general population is 0.5-3% (14–16). In a European population-based study, aneurysms were detected on screening MRI in about 1.8% of the adult population (17), while in a Norway population-based study the prevalence of intracranial aneurysms on MRA was 1.9% (18). These data suggest that the occurrence of UIA may be higher in the current cohort of patients with PCNSV. However, the high frequency of use of angiography in PCNSV may increase the likelihood of detection of small UIAs compared to non-invasive imaging studies. In a previous study, we observed that 16 (12.2%) of 131 patients with PCNSV had evidence of intracranial hemorrhage at or near the time of PCNSV diagnosis (19). Twelve patients had ICH, and 4 had SAH. Our cases 5 and 6 were included in that study. Despite the possible association between cerebral amyloid angiopathy (CAA) and intracranial hemorrhage (20), we did not observe in the previous study an increased frequency of CAA in patients with intracranial hemorrhage compared to those without. Seven of the 16 patients with intracranial hemorrhage in the previous study underwent brain biopsys and histologic evidence of vasculitis was observed in biopsy specimens from 4 patients. Necrotizing vasculitis was observed in 3 (75%) of these 4 positive biopsies. Similarly, in the present study our only biopsy-positive patient had evidence of a necrotizing vasculitis. Therefore, necrotizing vasculitis was the predominant histopathologic pattern in our patients presenting with intracranial hemorrhage or having evidence of UIA, being observed in 4/5 (80%) of biopsy-positive patients with these findings. Considering all biopsy-positive PCNSV patients, necrotizing vasculitis was the least frequent histologic pattern, present in only 14% of our PCNSV cohort (11, 21). The destructive vasculitic process with fibrinoid necrosis may cause severe vessel wall weakening, thus predisposing to blood vessel rupture and aneurysmal dilatation.

In this study, in most patients the co-occurring UIA was detected at PCNSV diagnosis, therefore we cannot be certain of a cause-effect relationship between PCNSV and UIA. Some of these aneurysms may have been present before the beginning of the vasculitic process. However, 2 patients (cases 3, and 10) developed aneurysms 8 and 10 months after PCNSV diagnosis or after the beginning of PCNSV clinical manifestations. In these 2 patients the first angiogram was negative for UIA and the development of UIA was associated with a worsening of the vasculitic process on imaging (appearance of new stenoses or worsening of the existing stenoses and/or appearance of new cerebral infarctions). High resolution contrast-enhanced black blood imaging was performed in one of these patients when the aneurysm was detected and showed segmental circumferential mural enhancement involving multiple vessels. In these 2 patients a relationship between the development of aneurysms and the vasculitic process was possible.

There was no change in the UIA in 10 patients during radiological follow-up for a median of 18.5 months. No patients developed new aneurysms and none had intracranial hemorrhage during the follow-up. All patients were treated with high dose prednisone and most of them also received additional immunodepressive therapy, particularly cyclophosphamide. In recent years different preclinical and clinical studies have established the important role of inflammation, particularly of interleukin-6 (IL-6), in potentiating intracranial aneurysm formation and rupture (22). Elevated IL-6 levels in the CSF and blood were correlated to subarachnoid hemorrhage severity and to intracranial aneurysm rupture (23–25). Some researchers speculated that the inhibition of inflammation and IL-6 may constitute a new therapeutic strategy for the prevention of intracranial aneurysm formation and rupture (22). In our patients, the immunosuppressive therapy started for the PCNSV could have conceivably prevented the rupture of aneurysms and the formation of new aneurysms, but smaller patient numbers prevent any definitive conclusions.

No clinical differences were observed between PCNSV patients with and without UIA except for a lower disability at last follow-up observed in patients with a UIA. The reasons for this finding are unclear. The increased medical surveillance during the follow-up period in the presence of a UIA may partly explain this result. This finding of favorable response to treatment and lower disability at last follow-up is similar to the previously described case series of PCNSV patients presenting with intracranial hemorrhage (19).

Adult PCNSV is a heterogeneous condition (1, 26), therefore it is important to identify patients with similar characteristics. We identified a homogeneous subgroup of PCNSV patients characterized by presenting with intracranial hemorrhage (19) or with the detection of UIA at diagnosis or during follow-up. They generally responded well to immunosuppressive treatment and had a good prognosis. These patients had a necrotizing vasculitis at brain biopsy indicating a common pathologic mechanism (27, 28).

This study has limitations, but also strengths. Its main limitations are the retrospective nature and the lack of histologic confirmation of diagnosis in two-thirds of the patients. Most of the patients were angiographically diagnosed, with long-term follow-up confirming the diagnosis of PCNSV. A potential referral bias may also limit the generalizability of the results. Strengths were the large number of unselected consecutive cases defined by uniform radiological and pathological criteria, the extensive detailed clinical data available, and the long-term follow up information.

## REFERENCES

1. Salvarani C, Brown RD Jr, Hunder GG. Adult primary central nervous system vasculitis. Lancet 2012; 380:767–777.

2. Salvarani C, Brown RD Jr, Hunder GG. Adult primary central nervous system vasculitis: an update. Curr Opin Rheumatol 2012; 24:46–52.

3. Salvarani C, Brown RD Jr, Christianson T, et al. An update of the Mayo Clinic cohort of patients with adult primary central nervous system vasculitis: description of 163 patients. Medicine 2015; 94:e738.

4. Salvarani C, Brown RD Jr, Calamia KT, et al. Primary central nervous system vasculitis: analysis of 101 patients. Ann Neurol 2007;62:442–451.

5. Salvarani C, Brown RD Jr, Calamia KT, et al. Primary CNS vasculitis with spinal cord involvement. Neurology 2008; 70:2394–2400

6. Calabrese LH, Mallek JA. Primary angiitis of the central nervous system. Report of 8 new cases, review of the literature, and proposal for diagnostic criteria. Medicine (Baltimore) 1988; 67:20–39.

7. Griffin J, Price DL, Davis L, McKhann GM. Granulomatous angiitis of the central nervous system with aneurysms on multiple cerebral arteries. Trans Am Neurol Assoc 1973; 98:145–8.

8. Yoong MF, Blumbergs PC, North JB. Primary (granulomatous) angiitis of the central nervous system with multiple aneurysms of spinal arteries. Case report. J Neurosurg 1993; 79:603–7.

9. Nishikawa M, Sakamoto H, Katsuyama J, et al. Multiple appearing and vanishing aneurysms: primary angiitis of the central nervous system. Case report. J Neurosurg 1998; 88:133–7.

10. Wen J, Ye S, Yang B, Liu X, Chen J. Multiple recurrent aneurysms with angiitis of the central nervous system in a girl: A case report. Medicine (Baltimore) 2022;101(51):e32415.

11. Salvarani C, Brown RD Jr, Christianson TJH, et al. Long-term remission, relapses and maintenance therapy in adult primary central nervous system vasculitis: a single-center 35-year experience. Autoimmun Rev 2020;19(4):102497.

12. Sulter G, Steen C, DeKeyser J. Use of the Barthel index and modified Rankin Scale in acute stroke trials. Stroke 1999; 30:1538–1541.

13. UCAS Japan Investigators; Morita A, Kirino T, Hashi K, et al. The natural course of unruptured cerebral aneurysms in a Japanese cohort. N Engl J Med 2012; 366:2474–82.

14. Brown RD Jr, Broderick JP. Unruptured intracranial aneurysms: epidemiology, natural history, management options, and familial screening. Lancet Neurol 2014; 13:393–404

15. Winn HR, Jane JA Sr, Taylor J, Kaiser D, Britz GW. Prevalence of asymptomatic incidental aneurysms: review of 4568 arteriograms. Stroke 1983; 14: 121.

16. Atkinson JL, Sundt TM Jr, Houser OW, Whisnant JP. Angiographic frequency of anterior circulation intracranial aneurysms. J Neurosurg 1989; 70: 551–55.

17. Vernooij MW, Ikram MA, Tanghe HL, et al. Incidental findings on brain MRI in the general population. N Engl J Med 2007; 357: 1821–28.

18. Müller TB, Sandvei MS, Kvistad KA, et al. Unruptured intracranial aneurysms in the Norwegian Nord-Trøndelag Health Study (HUNT): risk of rupture calculated from data in a population-based cohort study. Neurosurgery 2013; 73: 256–61.

19. Salvarani C, Brown RD Jr, Calamia KT, et al. Primary central nervous system vasculitis presenting with intracranial hemorrhage. Arthritis Rheum 2011; 63:3598–606.

20. Salvarani C, Hunder GG, Morris JM, Brown RD Jr, Christianson T, Giannini C. Aβ-related angiitis: comparison with CAA without inflammation and primary CNS vasculitis. Neurology 2013; 81:1596–603.

21. Giannini C, Salvarani C, Hunder G, Brown RD. Primary central nervous system vasculitis: pathology and mechanisms. Acta Neuropathol 2012;123:759–72.

22. Monsour M, Croci DM, Grüter BE, Taussky P, Marbacher S, Agazzi S. Cerebral Aneurysm and Interleukin-6: a Key Player in Aneurysm Generation and Rupture or Just One of the Multiple Factors? Transl Stroke Res. 2022 Aug 31. doi: 10.1007/s12975-022-01079-4. Online ahead of print.

23. Yao Y, Fang X, Yuan J, et al. Interleukin-6 in cerebrospinal fluid small extracellular vesicles as a potential biomarker for prognosis of aneurysmal subarachnoid haemorrhage. Neuropsychiatr Dis Treat 2021;17:1423–31.

24. Niwa A, Osuka K, Nakura T, Matsuo N, Watabe T, Takayasu M. Interleukin-6, MCP-1, IP-10, and MIG are sequentially expressed in cerebrospinal fluid after subarachnoid hemorrhage. J Neuroinflammation 2016;13(1):217.

25. Kao HW, Lee KW, Kuo CL, et al. Interleukin-6 as a prognostic biomarker in ruptured intracranial aneurysms. PLoS ONE 2015;10(7):e0132115

26. Hunder GG, Salvarani C, Brown RD Jr. Primary central nervous system vasculitis: Is it a single disease? Ann Neurol 2010; 68:573–4.

27. Miller DV, Salvarani C, Hunder GG, et al. Biopsy findings in primary angiitis of the central nervous system. Am J Surg Pathol 2009; 33:35–43.

28. Giannini C, Salvarani C, Hunder G, Brown RD. Primary central nervous system vasculitis: pathology and mechanisms. Acta Neuropathol 2012;123:759–72.

